# Age-dependent effects of adiposity on asthma risk during childhood and adulthood: a lifecourse Mendelian randomization study

**DOI:** 10.1101/2023.08.08.23293842

**Authors:** Helena Urquijo, Genevieve M Leyden, George Davey Smith, Tom G Richardson

**Affiliations:** MRC Integrative Epidemiology Unit (IEU), Population Health Sciences, Bristol Medical School, University of Bristol, Oakfield House, Oakfield Grove, Bristol, BS8 2BN, United Kingdom

**Keywords:** Adiposity, Childhood asthma, Mendelian randomization, Lifecourse epidemiology

## Abstract

**Background:** Separating the direct and long-term consequences of childhood lifestyle factors on asthma risk can be exceptionally challenging in epidemiology given that cases are typically diagnosed at various timepoints throughout the lifecourse.

**Methods:** In this study, we used human genetic data to evaluate the effects of childhood and adulthood adiposity on risk of pediatric (n=13,962 cases) and adult-onset asthma (n=26,582 cases) with a common set of controls (n=300,671) using a technique known as lifecourse Mendelian randomization.

**Findings:** We found that childhood adiposity increases risk of pediatric asthma (OR=1.20, 95% CI=1.03 to 1.37, P=0.03), whereas there was weak evidence that it has a long-term influence on adult-onset asthma (OR=1.05, 95% CI=0.93 to 1.17, P=0.39). Conversely, there was strong evidence that adulthood adiposity increases asthma risk in midlife using our lifecourse approach (OR=1.37, 95% CI=1.28 to 1.46, P=7×10^−12^).

**Interpretation:** These findings suggest that adiposity in childhood and adulthood are independent risk factors for asthma at each of their corresponding timepoints in the lifecourse. This inference would not be possible without the application of human genetic data, emphasizing the value of this approach in uncovering risk factors that begin to exert their influence on disease at early stages in life.

**Funding:** The Medical Research Council and the British Heart Foundation.

## Introduction

Pediatric asthma is the most common type of chronic disease among children which substantially contributes to the increasing global prevalence of asthma anticipated to affect 400 million individuals by 2025. One of the postulated explanations for this rise in cases is the escalating prevalence of childhood obesity, which has been reported to increase asthma risk based on findings from multiple study designs(1). However, establishing robust evidence of causal effects between risk factors and disease is notoriously challenging in a conventional epidemiological setting due to the influence of confounding factors and reverse causation. This is the motivation behind Mendelian randomization (MR), a causal inference technique which harnesses genetic variants quasi-randomly dispersed throughout a population to more reliably separate causal from nearly correlated risk factors (2, 3). That said, MR studies are often interpreted as ‘lifelong’ effects which presents a limitation in terms of interpreting causal relationships which may vary over the lifecourse, such as the effect of childhood adiposity on risk of asthma onset during childhood and later life.

We recently extended the principles of MR to investigate causal relationships which may vary throughout the lifecourse using genetic variants with time-varying effects(4, 5). For example, this approach previously found evidence that being overweight in childhood directly increases risk of type 1 diabetes, whereas the influence of adiposity on type 2 diabetes risk is largely attributed to a long-term and sustained effect of remaining overweight for many years over the lifecourse(6). In this current study, we sought to evaluate the effects of childhood and adult adiposity on asthma risk based on cases with recorded onset at two separate stages in life using data from the UK Biobank study (UKB)(7).

## Materials and Methods

### Instruments for lifecourse Mendelian randomization

Derivation of genetic instruments for childhood and adult adiposity have been described in detail previously(4). In brief, genome-wide association studies (GWAS) were conducted on 453,169 UKB participants who reported whether they were ‘thinner’, ‘about average’ or ‘plumper’ at age 10 years compared to the average. GWAS on the same sample were conducted based on their measured adult body mass index (mean age: 56.5 years old) which was categorised into the same proportions as the childhood adiposity variable for comparative purposes. These instrument sets have been extensively validated in three independent cohorts; the Avon Longitudinal Study of Parents and Children (ALSPAC) (4), the Young Finns Study (8) and the Trøndelag Health (HUNT) study (9)). Furthermore, a recent study has found that the childhood genetic instruments much more strongly influence DXA-derived total fat mass during childhood derived compared to lean mass (10).

### Pediatric- and adult-onset asthma datasets

Data on childhood-onset asthma were obtained from a previously conducted GWAS of 13,962 asthma cases diagnosed by a doctor before or at 19 years of age and 300,671 controls in UKB(7). The same study analysed 26,582 adult-onset asthma cases diagnosed between the ages of 20 to 60 years against the same set on controls (referred to as ‘adult-onset asthma’). The authors concluded that childhood-onset asthma has a partly distinct genetic architecture compared to asthma that first develops in later life, highlight differences in the pathophysiology between these disease subtypes(7).

### Ethics statement

All individual participant data used in this study were obtained from the UK Biobank (UKB) study, who have obtained ethics approval from the Research Ethics Committee (REC; approval number: 11/NW/0382) and informed consent from all participants enrolled in UKB.

### Statistical analysis

We firstly conducted univariable MR to estimate the total effects of childhood adiposity on asthma risk using data from the childhood and adulthood timepoints in turn. These estimates were derived using the inverse variance weighted (IVW) method, which takes SNP-outcome estimates and regresses them on the SNP-exposure associations (11). The weighted median (12) and MR-Egger (13) were used to assess the robustness of the IVW estimates to horizontal pleiotropy, whereby genetic variants influence multiple traits or disease outcomes via independent biological pathways (13). Multivariable MR, an extension of MR that employs multiple genetic variants associated with multiple measured risk factors, was used to calculate the direct and indirect effects of childhood and adulthood adiposity, simultaneously, on asthma risk at both timepoints (14, 15). Finally, to evaluate the robustness of our estimates to overfitting bias induced by overlapping samples between our exposures and outcomes, we applied a jackknife resampling approach to estimate effects (as described previously (16)). Analyses were conducted using the ‘TwoSampleMR’ R package. Forest plots in this paper were generated using the R package ‘ggplot2’ (17).

## Results

Firstly, we found strong evidence that childhood adiposity increases risk of childhood-onset asthma based on the inverse variance weighted (IVW) method (Odds Ratio (OR)=1.20, 95% Confidence Interval (CI)=1.03 to 1.37, P=0.03). Similar effect estimates using alternative MR approaches supported these findings (Figure 1A). In contrast, there was weak evidence that genetically predicted adulthood adiposity has an effect on childhood-onset asthma risk (e.g. IVW OR =0.97, 95% CI=0.84 to 1.10, P=0.68), providing a proof of concept for lifecourse MR given that in this instance the analysed outcome occurs earlier in life compared to the measured risk factor. Simultaneously estimating the effects of childhood and adult adiposity on childhood-onset asthma further suggested that being overweight in early life has a direct consequence on asthma risk at this early stage in the lifecourse (OR=1.36, 95% CI=1.14 to 1.57, P=0.005).

**Figure 1.**
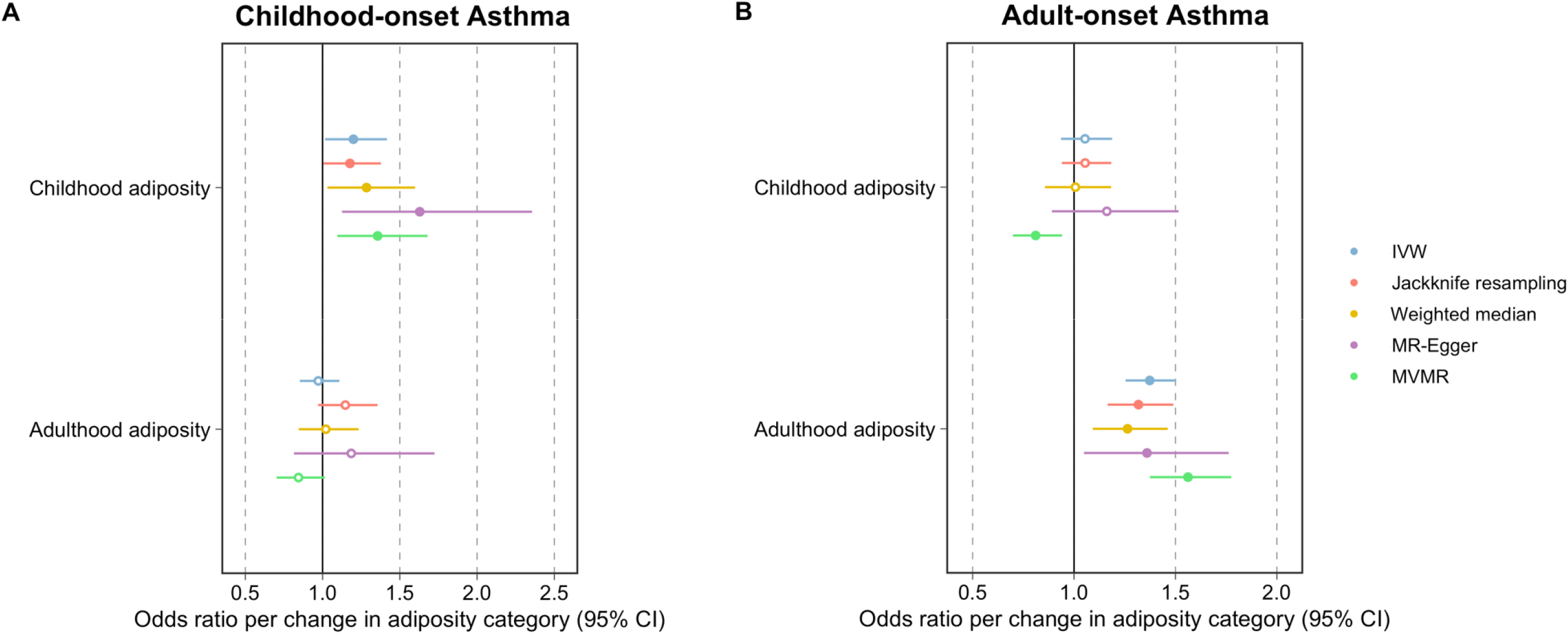
Forest plots illustrating lifecourse Mendelian randomization estimates of childhood and adulthood adiposity on A) childhood-onset and B) adult-onset asthma risk. IVW = inverse variance weighted method, MVMR = multivariable Mendelian randomization estimates

Conversely, using data on adulthood-onset asthma provided weak evidence that childhood adiposity has an effect on asthma risk in midlife (e.g. IVW OR=1.05 95% CI=0.93 to 1.17, P=0.39), whereas there was strong evidence that adulthood adiposity has an effect on asthma risk during adulthood (e.g. IVW OR =1.37, 95% CI=1.28 to 1.46, P=7×10^−12^). This finding was supported by multiple MR methods, as well as by estimates derived in a multivariable setting (OR =1.56, 95% CI=1.44 to 1.69, P=9×10^−13^) whilst simultaneously estimating the effect of childhood adiposity (Figure 1B). Estimates derived using jackknife resampling similarly provided concordance evidence of this effect, suggesting that overlapping samples in this study are unlikely to have substantially biased results.

## Discussion

Our findings provide compelling evidence that adiposity in childhood and adulthood are independent risk factors for asthma risk at each of their corresponding timepoints in the lifecourse. A key strength of our lifecourse MR approach is that it is capable of evaluating robust evidence of the direct effects of early life exposures, which would be extremely difficult to undertake without the use of human genetics, given the influence of confounding factors throughout the lifecourse. A proof of principle for this technique is the very limited evidence found in this study that adulthood adiposity has an effect on childhood-onset asthma risk, which is to be expected given that this outcome occurs at earlier stage in the lifecourse than the exposure.

Applying this approach herein provides evidence that a specific causal effect of adiposity exists during early life on risk of childhood-onset asthma risk, which therefore may develop into a long-life (and potentially immutable) disease. These results therefore support the hypothesis that one of the possible contributing factors for the rise in prevalence of childhood-onset asthma is the increasing numbers of individuals living with childhood obesity(18). This further emphasises the importance of introducing preventative strategies to help address the childhood obesity epidemic. Our findings suggest that this will help improve the quality of life for patients living with early-onset asthma as well as the many years over the lifecourse that this disease contributes to worldwide healthcare burdens.

## Data Availability

Data used in this study is based of previously published summary-level datasets.

## Author contributions

HU, GML and TGR conducted statistical analyses. All authors contributed to the design of the study and wrote the manuscript. All authors read and approved the final version of the manuscript.

## Materials and correspondence

This publication is the work of the authors and TGR will serve as guarantor for the contents of this paper.

## Acknowledgements

This research was conducted at the NIHR Biomedical Research Centre at the University Hospitals Bristol NHS Foundation Trust and the University of Bristol. The views expressed in this publication are those of the author(s) and not necessarily those of the NHS, the National Institute for Health Research or the Department of Health.

